# Benchmarking commercial healthcare claims data

**DOI:** 10.1101/2024.08.19.24312249

**Authors:** Alex Dahlen, Yaowei Deng, Vivek Charu

**Author notes:** Authors for correspondence: Alex Dahlen, PhD, Biostatistical Collaboration and Consultation Core (BC3), Department of Biostatistics, School of Global Public Health, New York University, Vivek Charu, MD PhD, Quantitative Sciences Unit, Departments of Medicine and Pathology, Stanford University School of Medicine.

## Abstract

**Importance:** Commercial healthcare claims datasets represent a sample of the US population that is biased along socioeconomic/demographic lines; depending on the target population of interest, results derived from these datasets may not generalize. Rigorous comparisons of claims-derived results to ground-truth data that quantify this bias are lacking.

**Objectives:** (1) To quantify the extent and variation of the bias associated with commercial healthcare claims data with respect to different target populations; (2) To evaluate how socioeconomic/demographic factors may explain the magnitude of the bias.

**Design:** This is a retrospective observational study. Healthcare claims data come from the Merative™ MarketScan® Commercial Database; reference data for comparison come from the State Inpatient Databases (SID) and the US Census. We considered three target populations, aged 18-64 years: (1) all Americans; (2) Americans with health insurance; (3) Americans with commercial health insurance.

**Participants:** We analyzed inpatient discharge records of patients aged 18-64 years, occurring between 01/01/2019 to 12/31/2019 in five states: California, Iowa, Maryland, Massachusetts, and New Jersey.

**Outcomes:** We estimated rates of the 250 most common inpatient procedures, using claims data and using reference data for each target population, and we compared the two estimates.

**Results:** The average rate of inpatient discharges per 100 person-years was 5.39 in the claims data (95% CI: [5.37, 5.40]) and 7.003 (95% CI: [7.002, 7.004]) in the reference data for all Americans, corresponding to a 23.1% underestimate from claims. We found large variation in the extent of relative bias across inpatient procedures, including 22.8% of procedures that were underestimated by more than a factor of 2. There was a significant relationship between socioeconomic/demographic factors and the magnitude of bias: procedures that disproportionately occur in disadvantaged neighborhoods were more underestimated in claims data (*R*^2^ = 51.6%, p < 0.001). When the target population was restricted to commercially insured Americans, the bias decreased substantially (3.2% of procedures were biased by more than factor of 2), but some variation across procedures remained.

**Conclusions and relevance:** Naïve use of healthcare claims data to derive estimates for the underlying US population can be severely biased. The extent of bias is at least partially explained by neighborhood-level socioeconomic factors.

## Introduction

Commercial healthcare claims databases are among the largest patient-level data sets ever assembled, offering enormous potential for clinical research. They are increasingly being used in the context of disease monitoring and comparative effectiveness research^1–7^. Despite their widespread use, commercial healthcare claims databases have not been rigorously vetted against ground-truth data.

These datasets are assembled by aggregating data from commercial insurers and, as such, they represent a non-random sample of the underlying population. In prior work, we have demonstrated that inclusion in large commercial healthcare claims databases varies spatially and is systematically biased along socioeconomic and demographic lines, compared to the overall US population: patients who are old, White, wealthy, or college-educated are over-sampled relative to other populations^8^. Statistical inferences derived from non-random samples are susceptible to external validity bias and may fail to generalize to the underlying target population of interest^9^. In the context of claims data, this bias arises when the same social determinants of health associated with inclusion in the datasets also affect the health outcomes of interest (e.g. disease burden, healthcare access, or treatment effectiveness).

Relatively few studies have attempted to quantify external validity bias in claims-derived results, in part because ground-truth data on the outcome of interest is often unavailable. In the setting of infectious diseases, with regional surveillance data serving as the ground-truth, prior work has demonstrated that claims-derived incidence of measles, mumps and varicella are dramatically overestimated^10^; and, though claims-derived *incidence rates* of influenza were inaccurate, claims-derived *disease patterns* were representative^11,12^. The RCT-Duplicate study has made direct comparisons between treatment effect estimates derived from 32 randomized controlled clinical trials (RCTs) and those estimated from claims data, demonstrating that claims-derived inferences were generally comparable in a highly selected and non-representative set of RCTs^13^. Taken together, these prior studies highlight challenges in quantifying external validity bias in claims-derived results, and demonstrate that the extent of bias depends on the disease context and outcome of interest.

Here, we present the most detailed empirical analysis of external validity bias in healthcare claims data to date, focusing on the rates of inpatient procedures, for which a unique ground-truth dataset exists. We quantify the extent and variation in external validity bias across a comprehensive set of inpatient procedures, and evaluate how social determinants of health explain the magnitude of bias.

## Methods

This cross-sectional study was approved by the Stanford University institutional review board IRB 40974). Reporting followed the STROBE reporting guideline.

### Data

For healthcare claims data, we used the Merative™ MarketScan® Commercial Database (MarketScan). It includes the health service data for approximately 250 million privately insured employees and dependents in the United States with primary healthcare coverage through fee-for-service, point-of-service or capitated health plans. All enrollment records and inpatient, outpatient, ancillary and drug claims are collected. Patient-level demographic information has been de-identified except for age, gender, and the state and Metropolitan Statistical Area of residence. The dataset’s digital object identifier (DOI) is: 10.57761/n5v8-0v21.

Ground-truth data were derived from the State Inpatient Databases (SIDs), which are part of a family of databases maintained by the Healthcare Cost and Utilization Project (HCUP), and sponsored by the Agency for Healthcare Research and Quality. The SIDs provide all inpatient discharges from non-federal acute care hospitals; they include information on patient demographics, primary and secondary diagnosis and procedures codes, health insurance status/type, hospital charges and length of stay. Because the SIDs capture more than 97% of all hospital discharges in each state, we considered data derived from the SIDs as ground-truth. Patient/discharge-level SID data includes: age, gender, insurance type and the patient’s zip code. State populations and demographic information to characterize the overall cohort were extracted from the 2019 American Community Survey (ACS) 5-year census data.

We used a convenience sample of SIDs from California, Iowa, Michigan, Maryland and New Jersey, and analyzed all inpatient discharges that occurred during the period 01/01/2019 to 12/31/2019 for patients in the age range 18-64 (at the time of discharge). Likewise, for the MarketScan data, we analyzed all inpatient discharges that occurred during the same period (01/01/2019 to 12/31/2019), for patients with the same age range (18-64), residing in the same states (California, Iowa, Michigan, Maryland and New Jersey). Both SID and MarketScan inpatient data exclude ambulatory and outpatient procedures. As described in the supplementary material, we restricted both datasets to acute-care facilities and to procedures occurring therein.

### Outcomes of interest

We compared the rates of inpatient procedures between claims data and the ground truth data. Inpatient procedures were classified using the Clinical Classification Software Refined (CCRS) classification of ICD-10-PCS codes^14^, and we studied the 250 most common inpatient procedures in the SID dataset, after excluding a small subset of procedures with extreme distributions of coding by state (see supplementary material). A list of the procedure codes used to identify each inpatient procedure is provided in **Table S1**. We chose to study inpatient procedures because, while diagnosis codes are often carried over across multiple encounters with the healthcare system, procedure codes reflect a procedure performed and billed for at a specific encounter.

### Characterizing social determinants of health (SDOH)

We use two strategies to measure aggregate neighborhood-level SDOH: our primary measure was zip code-level National Deprivation Index (NDI) which is a single metric of deprivation defined and maintained by the National Cancer Institute^15^; 13 socioeconomic indicators are extracted from the 2017 5-year census and combined into a single measure of deprivation. We used population averages to roll the score up from the census tract-level to the zip code-level. As a sensitivity analysis, we used principal component analysis (PCA) to reduce 25 socioeconomic and demographic indicators extracted from the 2019 census to a single metric of socioeconomic status (see supplementary methods).

### Statistical analysis

The goal of our analysis was to (1) quantify the potential bias in estimates of the prevalence of a large number of inpatient procedures derived from MarketScan and (2) characterize factors associated with the size of the bias.

#### Quantifying the bias

We estimated the rate of each procedure in the claims data and the SID data separately. For the claims data, rates were estimated by dividing the number of inpatient discharges with the appropriate ICD-10-PCS codes by the total number of patient-years of coverage in the dataset, across the entire cohort. For the reference data, rates were estimated by dividing the number of discharges with the appropriate ICD-10-PCS code in the SID data by the 2019 population estimate for our cohort as derived from ACS data.

We defined the relative bias for each procedure as the ratio of the claims-derived rate divided by the ground truth: relative bias = rate derived from claims data/rate derived from reference data. A relative bias of 1 indicates that claims-derived estimates align with ground truth data (no bias); values less than 1 indicate that the claims data underestimate the rate, and values greater than 1 indicate that claims data overestimate it. We computed 95% Poisson confidence intervals for both rate estimates, and errors were propagated to the relative bias using the delta method on the log of the ratio.

#### Characterizing factors associated with the bias

We hypothesized that the magnitude of the relative bias for a given procedure would depend strongly on the social determinants of health of the patient population that undergoes each procedure. In particular, since people who are old, White, wealthy, or college-educated are over-sampled in the claims data^8^, we hypothesized that procedures that are disproportionately performed on those demographic groups will tend to be overestimated, and vice versa.

For each procedure, we evaluated the strength of the association between the procedure and SDOH using a zip code-level Poisson regression of the form:

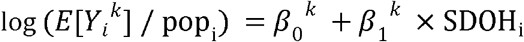

where *k* indexes the procedures, and *i* indexes zip codes. For each zip code *i, Y_i_*^*k*^ is the count of the number procedures, pop*_i_* is the population, and SDOHi is a zip code-level proxy metric of aggregate SDOH_i_. In our primary analysis, we use NDI as this proxy metric, and in a sensitivity analysis, we use the SES metric we defined by PCA. For each procedure *k, β_1_*^*k*^ measures the strength of the association between SDOH and the procedure rate (across zip codes). Larger positive values of *β_1_*^*k*^ indicate that the procedure disproportionately occurs in zip codes with higher levels of the SDOH metric, and vice versa.

Finally, we used a log-linear regression to quantify the association between the relative bias for each procedure and *β_1_*^*k*^. A significant positive association between these quantities would indicate that the procedures that are most underestimated are those that tend to be performed in zip codes with lower NDI. We estimated an *R*^2^ value to quantify the fraction of the variation in relative bias explained by the association between NDI and the rate of the procedure.

#### Quantifying the bias for different target populations

Lastly, we are interested in understanding how the relative bias changes as we change the target population of interest. For our primary analysis described above, the target population was taken to be all Americans (aged 18-64 in 2019), but we also considered two other possible target populations: *insured* Americans and *commercially insured* Americans. As the target population was restricted to more closely resemble the population represented in the claims dataset, we expected to see the overall bias decrease, but we were still interested in quantifying how much variation in the bias still existed. To compute reference estimates for these two additional target populations, we insurance type for both the SID data and the ACS Census population estimates to filter down to the relevant group.

### Reproducibility

Full details of the statistical methods are provided in supplementary material. The analysis was conducted in Python version 3.8.5, and the code has been shared publicly at https://github.com/alex-dahlen/ClaimsDataBenchmarking.

## Results

We identified ∼2.95 million hospital discharges among our cohort from January 1 to December 31, 2019, in the State Inpatient Databases (SIDs) (**Table 1**). For the same time period and age group, ∼660k hospital discharges were identified in the claims dataset. Demographic information for the underlying populations are provided in **Table 1**.

**Table 1.** A comparison of the two cohorts: the claims-data cohort (MarketScan) and the ground-truth reference cohort (ACS 5-year data Census data). For both cohorts, inclusion criteria are people aged 18-64, in the year 2019, residing in one of 5 states: CA, IA, MD, MI, and NJ. For the claims data, we record the number of days of coverage during 2019 for each member of the cohort; for the reference group, we assume members remain in the cohort for the entire year. MarketScan provides limited demographic data on its members (just age and sex); the additional demographic details about the reference cohort are derived from the ACS 5-year 2019 Census. Note: some members in the claims-data cohort had coverage across two or more states in 2019, so the percentages slightly exceed 100%. *The number of inpatient discharges was derived from the State Inpatient Databases (SID).

|  | **Claims Data (MarketScan)**<br>2019, ages 18-64<br>5 states (CA, IA, MD, MI, NJ) | **Ground truth (Census)**<br>2019, ages 18-64<br>5 states (CA, IA, MD, MI, NJ) |
| --- | --- | --- |
| n | 2,793,302 | 42,062,788 |
| Days of coverage |  |  |
| Mean (SD) | 303.9 (103.7) | 365 (0) |
| Median [IQR] | 365 [273-365] | 365 [365-365] |
| Inpatient visits |  |  |
| Number of discharges | 658,950 | 2,945,667* |
| Inpatient discharges / 100 patient-years | 5.39 | 7.00 |
| Female (%) | 1,403,619 (50.2%) | 21,014,821 (50.0%) |
| State (%) |  |  |
| CA | 1,192,060 (42.7%) | 24,775,310 (58.9%) |
| IA | 97,481 (3.5%) | 1,885,249 (4.5%) |
| MD | 276,515 (9.9%) | 3,774,488 (9.0%) |
| MI | 705,132 (25.2%) | 6,121,044 (14.6%) |
| NJ | 524,596 (18.8%) | 5,506,697 (13.1%) |
| Age, n (%) |  |  |
| 18-34 | 1,026,345 (36.7%) | 11,320,834 (26.9%) |
| 35-49 | 872,083 (31.2%) | 13,320,352 (31.7%) |
| 50-64 | 894,874 (32.0%) | 17,421,602 (41.4%) |
| Health insurance, n (%) |  |  |
| Commercial insurance | 100% | 29,339,545 (72.3%) |
| Public (Medicaid) | 0 (0%) | 7,197,270 (17.7%) |
| Uninsured | 0 (0%) | 4,015,926 (9.9%) |
| Race / ethnicity, n (%) |  |  |
| Hispanic | Unknown | 19,256,925 (28.4%) |
| Non-Hispanic Asian | Unknown | 7,693,855 (11.4%) |
| Non-Hispanic Black | Unknown | 6,367,232 (9.4%) |
| Non-Hispanic White | Unknown | 30,772,555 (45.5%) |
| Other | Unknown | 3,599,769 (5.3%) |
| Education, n (%) |  |  |
| Less than high school | Unknown | 6,241,773 (13.7%) |
| High school | Unknown | 10,728,771 (23.5%) |
| Some college | Unknown | 13,025,249 (28.5%) |
| College | Unknown | 9,595,841 (21.0%) |
| Graduate | Unknown | 6,071,561 (13.3%) |
| Household income, n (%) |  |  |
| < \$20k | Unknown | 3,030,399 (12.8%) |
| \$20k – \$40k | Unknown | 3,663,457 (15.5%) |
| \$40k – \$75k | Unknown | 5,494,523 (23.2%) |
| \$75k – \$125k | Unknown | 5,283,574 (22.3%) |
| \$125k – \$200k | Unknown | 3,626,380 (15.3%) |
| > \$200k | Unknown | 2,583,525 (10.9%) |

The overall estimated rate of all inpatient discharges was 7.003 [7.002, 7.004] per 100 person-years derived from the reference data, and 5.39 [5.37, 5.41] derived from claims data. This corresponds to a relative bias (claims / reference) of 0.769 (95% CI: [0.767, 0.771]), indicating that, on average, the claims data underestimated the rate of all inpatient discharges by 23.1%.

We found considerable variation in estimates of this relative bias across the most common 250 procedures. The ten most under- and overestimated procedures are shown in **Table 2**, and a forest plot of the relative bias all 250 procedures is shown in **Figure 1**. (See **Supplementary Excel File** for full results.) We found that 50.4% of procedures were under- or over-estimated by more than a factor of 1.5, and 22.8% by more than a factor of 2 (**Supp Table 1**).

**Table 2.** The top 10 most underestimated and the top 10 most overestimated procedures in our analysis. The rate of each procedure was estimated using claims data (MarketScan) and ground-truth reference data (SID and Census). The relative bias is determined by taking the ratio between the two estimates (claims / reference). Relative biases that are smaller than 1 correspond to cases where claims data has underestimated the rate and vice-versa.

| Procedure | Claims Data<br>rate / 100 patient-years | Reference Data<br>rate / 100 patient-years | Relative Bias<br>Claims / Reference |
| --- | --- | --- | --- |
| **Top 10 most overestimated procedures** |  |  |  |
| Knee arthroplasty | 0.1275 ± 0.0020 | 0.08759 ± 0.00009 | 1.456 [1.433, 1.479] |
| Gastro-jejunal bypass (including bariatric) | 0.0334 ± 0.0010 | 0.02336 ± 0.00005 | 1.429 [1.385, 1.473] |
| Prostatectomy | 0.0172 ± 0.0007 | 0.01259 ± 0.00003 | 1.368 [1.310, 1.427] |
| Hip arthroplasty | 0.1092 ± 0.0019 | 0.08595 ± 0.00009 | 1.271 [1.250, 1.293] |
| Prostate and seminal vesicle procedures (excluding prostatectomy) | 0.0086 ± 0.0005 | 0.00696 ± 0.00003 | 1.237 [1.165, 1.314] |
| Heart conduction mechanism procedures | 0.0107 ± 0.0006 | 0.00896 ± 0.00003 | 1.197 [1.134, 1.263] |
| Administration and transfusion of bone marrow, stem cells, pancreatic islet cells, and t-cells | 0.0064 ± 0.0005 | 0.00546 ± 0.00002 | 1.180 [1.101, 1.266] |
| Breast reconstruction | 0.0177 ± 0.0007 | 0.01518 ± 0.00004 | 1.169 [1.121, 1.219] |
| Colectomy | 0.0678 ± 0.0015 | 0.06058 ± 0.00007 | 1.120 [1.096, 1.144] |
| Lymph node excision (therapeutic) | 0.0138 ± 0.0007 | 0.01238 ± 0.00003 | 1.116 [1.064, 1.171] |
| **Top 10 most underestimated procedures** |  |  |  |
| Subcutaneous contraceptive implant | 0.0016 ± 0.0002 | 0.00903 ± 0.00003 | 0.172 [0.149, 0.198] |
| Cardiac stress tests | 0.0019 ± 0.0002 | 0.00939 ± 0.00003 | 0.198 [0.174, 0.226] |
| Hemodialysis | 0.0462 ± 0.0012 | 0.22782 ± 0.00015 | 0.203 [0.198, 0.208] |
| Transfusion of plasma | 0.0118 ± 0.0006 | 0.05375 ± 0.00007 | 0.220 [0.209, 0.232] |
| Cardiac chest compression | 0.0062 ± 0.0004 | 0.02704 ± 0.00005 | 0.230 [0.214, 0.247] |
| Arterial oxygen saturation monitoring | 0.0020 ± 0.0003 | 0.00751 ± 0.00003 | 0.271 [0.239, 0.307] |
| Administration of diagnostic substances, NEC | 0.0019 ± 0.0002 | 0.00692 ± 0.00003 | 0.276 [0.243, 0.314] |
| Peripheral arteriovenous fistula and shunt procedures | 0.0017 ± 0.0002 | 0.00617 ± 0.00002 | 0.278 [0.243, 0.319] |
| Finger and other upper extremity amputation | 0.0009 ± 0.0002 | 0.00318 ± 0.00002 | 0.280 [0.232, 0.338] |
| Peripheral arterial pressure monitoring | 0.0068 ± 0.0005 | 0.02380 ± 0.00005 | 0.286 [0.267, 0.306] |

**Fig 1.**
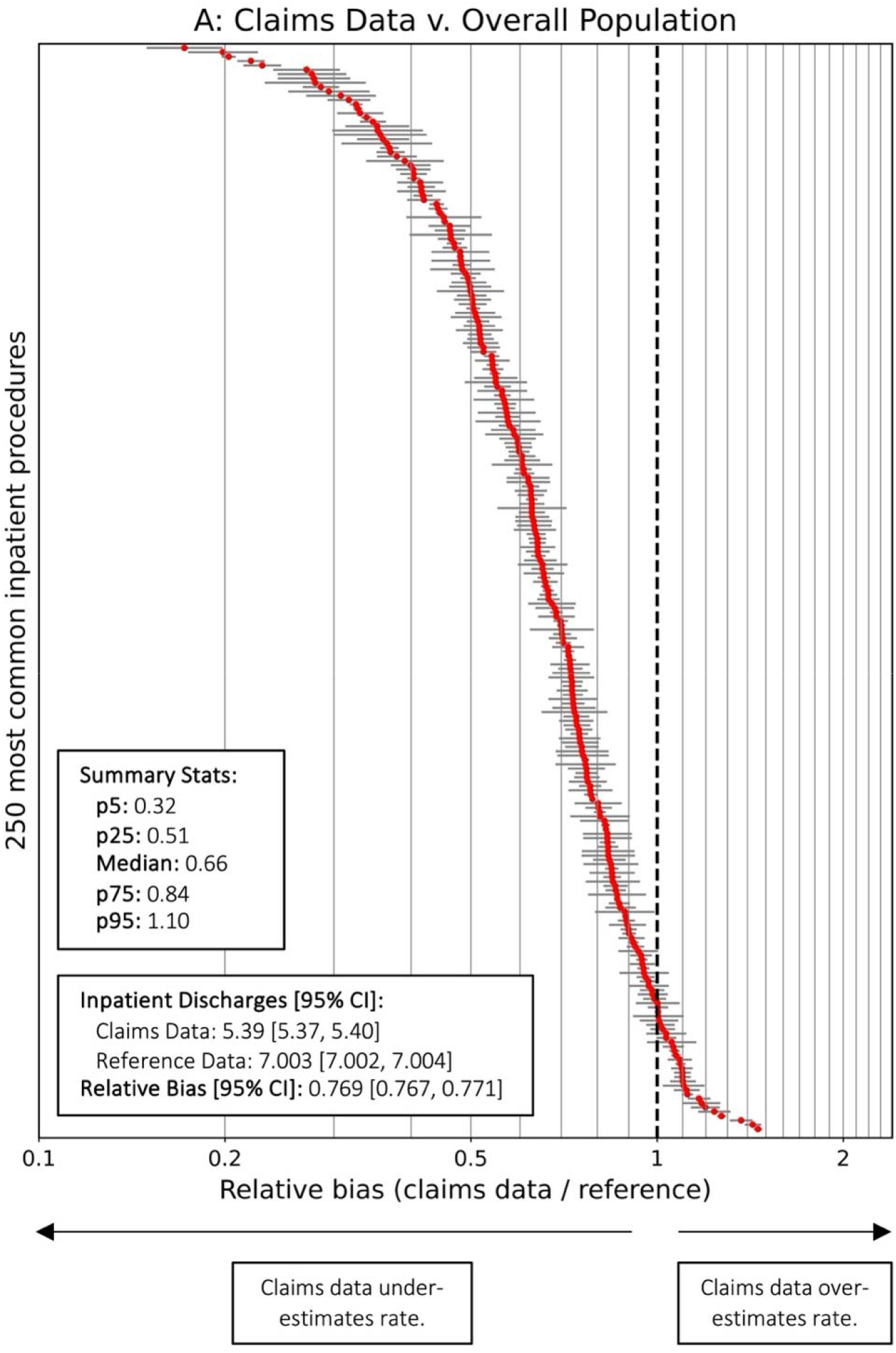
Forest plot of the relative bias for all 250 procedures. The overall estimated rate of all inpatient discharges was 5.4 / 100 person-years in the claims data compared to 7.0 /100 person-years in the reference data, corresponding to a relative bias of 0.77, or an underestimate of 23% (the reference target population is All Americans in this analysis). This forest plot shows the relative bias (and 95% confidence interval) for each of the 250 most common inpatient procedures, ordered by the magnitude of the bias. There is large variation in the extent of the bias across different procedures: 25% of procedures are underestimated by 50% of more, and another 5% of procedures are over-estimated by 10% or more.

**Fig 2.**
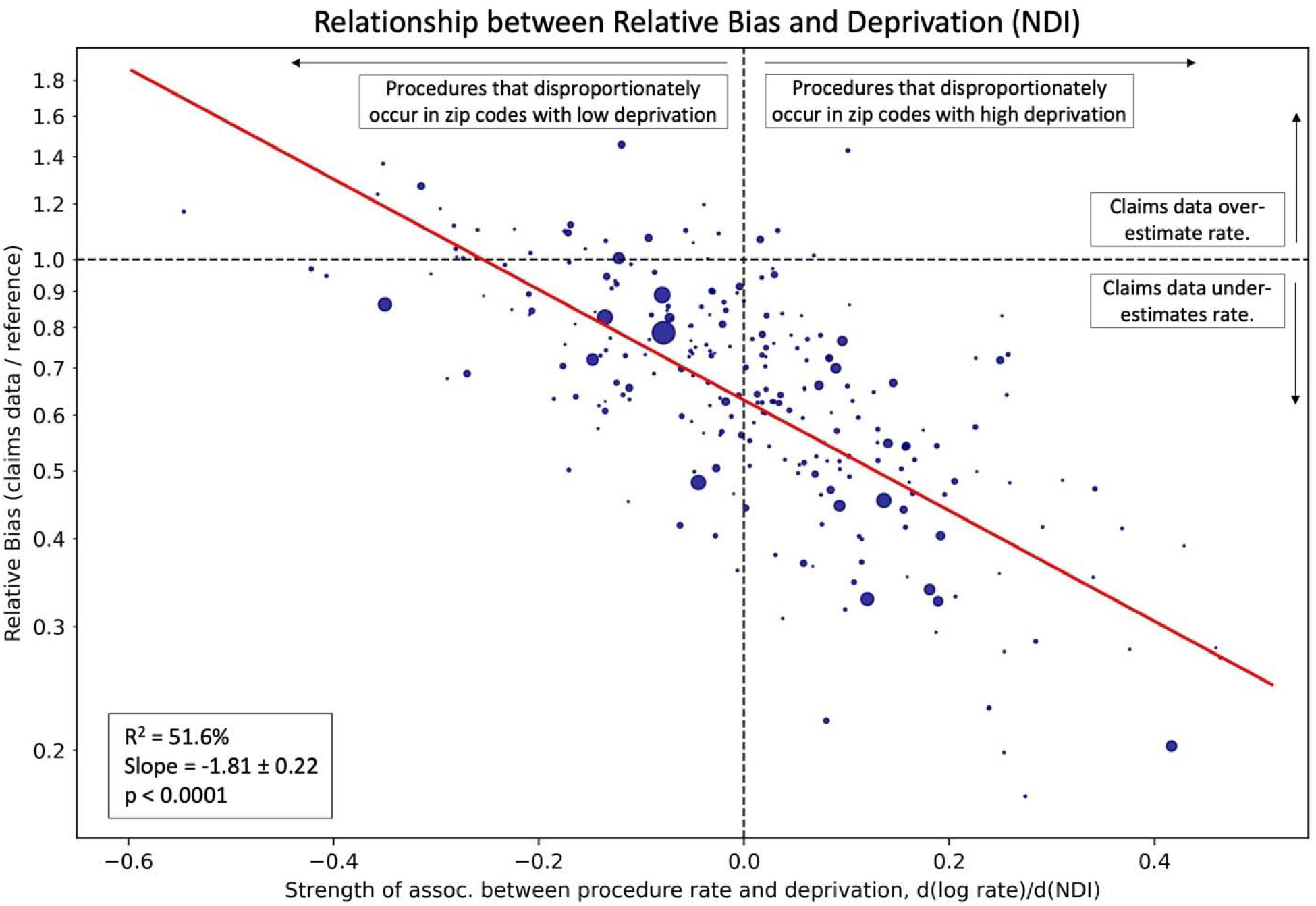
The relationship between the relative bias for each procedure and the association between the procedure rate and the neighborhood deprivation index (NDI). For each procedure, we computed the latter association by comparing zip code-level procedure rates with NDI; higher levels of this association indicate that the procedure is disproportionately performed in zip codes with high deprivation and vice versa. Examples of procedures with that are disproportionately performed in high deprivation areas are: hemodialysis, above knee amputation and arterial oxygen saturation monitoring. Examples of procedures that are disproportionately performed in low deprivation areas are: breast reconstruction, prostatectomy and hip replacement. We find that procedures disproportionately performed in high deprivation areas are far more likely to be underestimated and vice versa (p<0.0001). We obtained similar results when we defined an alternative measure of neighborhood socioeconomic status using 2019 census data, and the results of that sensitivity analysis are shown in **Supp. Fig. 5**.

Overestimated procedures included: knee replacements (overestimated by 46%), hip replacement (27%), gastric bypass (43%), and prostatectomy (37%); severely underestimated procedures included: hemodialysis (underestimated by 80%), cardiac stress tests (80%), subcutaneous contraceptive insertion (83%), and transfusion of plasma (78%).

We found a clear relationship between the relative bias for a given procedure, and the procedure’s association with social determinants of health, measured via the Neighborhood Deprivation Index (**Figure 3;** R^2^ = 51.6%, p<0.0001, slope = −1.81). Procedures that are disproportionately performed in neighborhoods with higher levels of deprivation (NDI) were significantly more likely to be underestimated, and vice versa. Some examples of procedures that were disproportionately performed in neighborhoods with high deprivation are: hemodialysis (underestimated by 80%), finger and other upper extremity amputation (underestimated by 72%), and arterial oxygen saturation monitoring (underestimated by 73%). Examples of procedures that were disproportionately performed in neighborhoods with low deprivation are: breast reconstruction (overestimated by 17%), prostatectomy (overestimated by 37%), and hip replacement (overestimated by 27%). As a sensitivity analysis, we performed a similar analysis with a socioeconomic status metric we defined using 2019 census data, and we found an even stronger association (R^2^ = 58.8%, p<0.0001, **Supp. Fig. 5**), perhaps the alternative metric is derived from data overlapping with the time period of our analysis, while the NDI metric is derived from 2017 data.

**Fig 3.**
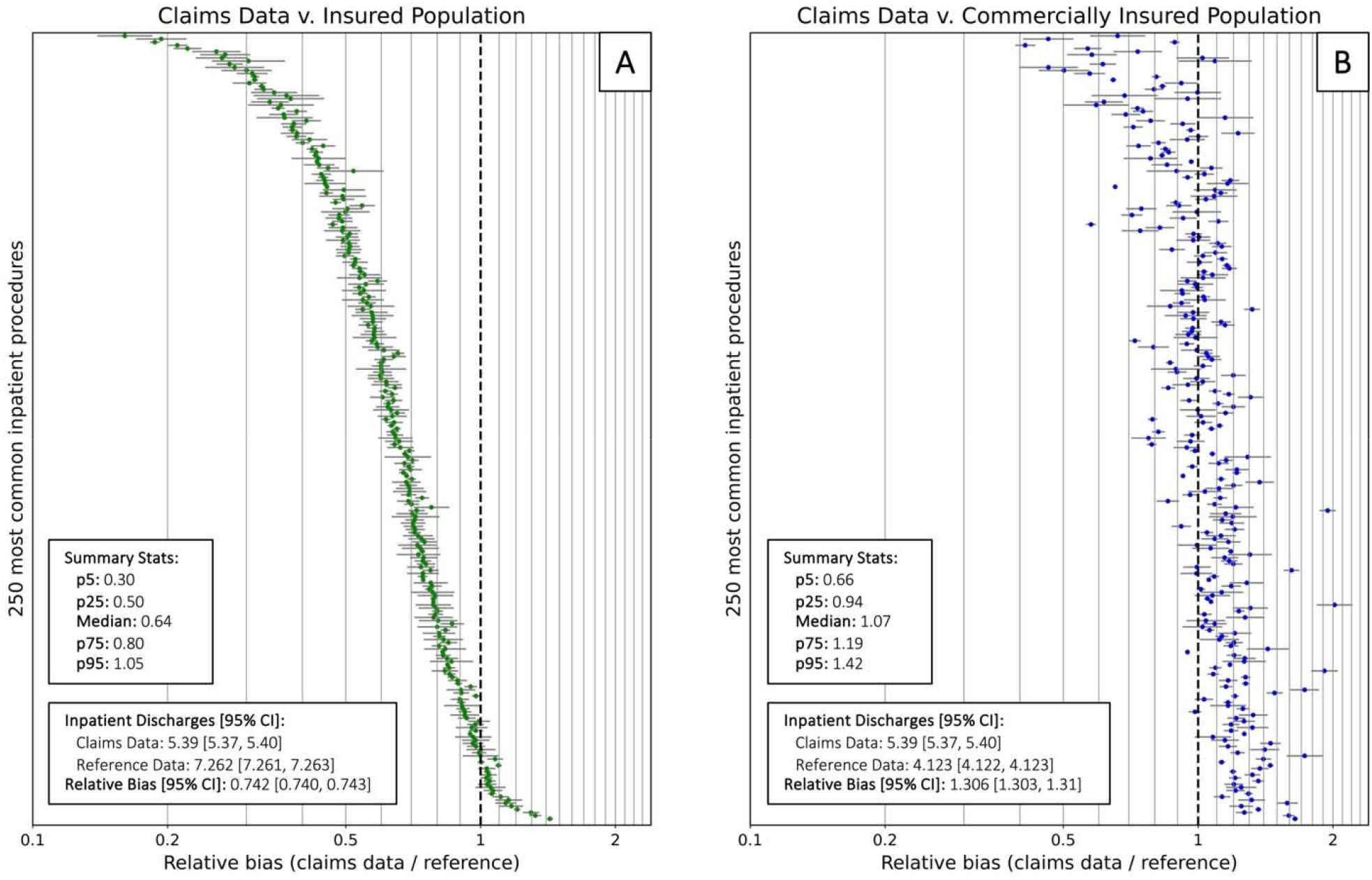
The impact of changing the target population. **Fig 3A** displays a forest plot of the relative bias for all 250 procedures, restricting the reference data to the insured population; **Fig 3B** displays a forest plot of the relative bias for all 250 procedures, restricting the reference data to the commercially insured population. (For all target populations, we focused on the subset that are aged 18-64 in the year 2019, and living in CA, IA, MD, MI, or NJ.). Procedures are ordered on the y-axis in the same way as they are in Fig. 1.

When we restricted the target population to those with any health insurance (**Figure 3A**), we found the overall pattern of bias was similar, likely because patients without health insurance account for less than 10% of total inpatient discharges in the reference data. However, when we further restricted the target population to those with *commercial* health insurance (**Figure 3B**), the pattern of bias changes. The overall rate of all inpatient discharges amongst the commercially insured in our reference cohort was 4.123 [4.122, 4.123] per 100 person-years; compared to this, the claims data estimate (5.39 [5.37, 5.41]) is a 30.6% overestimate. The relative bias for most procedures improved: while the interquartile range for the relative biases was [0.51, 0.84] when comparing to the overall population; it was [0.94, 1.19] when comparing to the commercially insured population, which is narrower and, notably, overlaps 1. Nonetheless, there is still some variation in the bias by procedure; 11.2% of procedures were under- or over-estimated by more than a factor of 1.5, and 3.2% by more than a factor of 2. When we re-evaluated the relationship between the relative bias and the and the strength of the association with SDOH for this new target population, we found a much attenuated association (R^2^ = 3.4%, p=0.005, slope = −0.26; **Supp. Figure 6**).

## Discussion

Healthcare claims data are increasingly being used to evaluate disease burden and quantify the effects of health policies and biomedical treatments on health outcomes. These datasets are a non-random sample of the US population. Prior work has shown that, compared to the US population, inclusion in large commercial healthcare claims databases is biased geographically and along socioeconomic and demographic lines^8^. When the outcome of interest is associated with these socioeconomic and demographic features, claims-derived inferences are susceptible to external validity bias. While methods have been developed to translate clinical trial results to different target populations^16^, these methods cannot be directly applied to claims data results, since claims datasets typically lack individual-level information on socio-economic/demographic indicators to readily allow for transport of effects to different populations of interest. Isolated studies have evaluated the bias for selected outcomes^10,11^, but they have been limited because of the difficulty of obtaining ground-truth reference data.

Here we report on the empirical bias in healthcare claims-derived estimates of the 250 most common inpatient procedures for US patients aged 18-64, using a unique ground-truth dataset of inpatient hospitalizations. We found that: (1) with respect to all Americans, commercial healthcare claims data underestimate the true incidence of overall inpatient visits by ∼27%, reflecting lower inpatient healthcare utilization among commercially insured patients; (2) the extent of the bias varies considerably across inpatient procedures, with 22.4% of procedures being under or overestimated by more a factor 2; (3) procedures that disproportionately occur in patients from low SES neighborhoods are the most severely underestimated and vice versa; (4) if healthcare claims data are compared to a restricted target population of commercially insured Americans, the magnitude of external validity bias is considerably attenuated, but there is still some variation in the bias across different procedures and it is at least partially explained by SDOH.

A strength of our study is the focus on inpatient procedures, with outcome definitions as similar as possible between the claims and ground-truth data. All studies that make use of claims data are susceptible to misclassification bias^17^, where errors diagnosing or coding for a disease occur. By focusing on two datasets with identical outcome definitions, we attribute the bias we have measured to non-random sampling.

Several limitations must be mentioned. First, we relied on a convenience sample of SID data, limited to 5 states. Although this sample of states was non-random, it reflects over 20% of the US population in year 2019, and covers a reasonable geographic distribution. Second, while the SID data are extensive, up to 5% of inpatient visits are not captured; nonetheless we believe this is as close to a ground-truth reference dataset we can achieve for this purpose. Third, because the SID data only provides region information at the zip code-level, our estimate of the strength of the association between a procedure and NDI was defined at the zip code-level. More granular regional information could improve this metric. Finally, we restricted this study to the bias associated with inferences of rates of inpatient procedures; many other types of inferences remain to be studied, including comparative effect sizes.

Despite these limitations, our analysis has several implications for studies analyzing healthcare claims data. Whenever the outcome (or treatment effect) of interest is either associated with or modified by SDOH, claims-derived results can be biased. In particular: (1) studies that seek to estimate disease prevalence/incidence rates and medication prescription rates are very likely to be biased with respect to the US population, since SDOH are known to be associated with disease burden and access to health care; (2) studies that seek to estimate treatment effects by using claims data to emulate an RCT will be biased whenever treatment (or access to treatment, or adherence to treatment) is related to SDOH; and (3) studies that seek to evaluate the impact of policy-level changes will be biased if the policy of interest has heterogenous effects across either insurance status or SDOH. To improve the transparency and reliability of studies using healthcare claims data, investigators should provide a first-principles argument for how SDOH might or might not moderate the outcome of interest. Additionally, whenever possible, studies should seek to replicate their findings in more than one claims dataset with different patterns of sampling (e.g. Medicaid claims).

Because of their large sample size, healthcare claims data offer enormous potential in research; characterizing and overcoming this selection bias is an essential first step to unlocking their potential.

## Data Availability

The dataset's digital object identifier (DOI) is: 10.57761/n5v8-0v21.

## Acknowledgements

Data for this project were accessed using the Stanford Center for Population Health Sciences Data Core. The PHS Data Core is supported by a National Institutes of Health National Center for Advancing Translational Science Clinical and Translational Science Award (UL1TR003142) and from Internal Stanford funding. The content is solely the responsibility of the authors and does not necessarily represent the official views of the NIH.

## Funding

This work was supported by KL2TR003143 (VC), P30DK116074 (VC). The funders had no role in study design, data collection and analysis, decision to publish, or preparation of the manuscript.

## Guarantor

AD and VC take full responsibility for the work as a whole, including the study design, access to data, and the decision to submit and publish the manuscript.

## Conflicts of interest

The authors have no conflicts of interest to disclose.

## Authors’ Contributions

AD: Research idea and study design, data analysis and interpretation, statistical analysis

YD: Data analysis and interpretation, statistical analysis

VC: Research idea and study design, data analysis and interpretation, statistical analysis

Each author contributed important intellectual content during manuscript drafting or revision and agrees to be personally accountable for the individual’s own contributions and to ensure that questions pertaining to the accuracy or integrity of any portion of the work, even one in which the author was not directly involved, are appropriately investigated and resolved, including with documentation in the literature if appropriate.

## Notes

### Competing Interest Statement

The authors have declared no competing interest.

### Funding Statement

This study was funded by KL2TR003143.

### Author Declarations

IRB of Stanford University waived ethical approval for this work. This is an analysis of de-identified datasets, covered by Stanford University IRB 40974.

