## Supplementary Materials for "Benchmarking commercial healthcare claims data"

**Detailed Statistical Methods**

This Supplement provides the full statistical details of the three steps of our analysis. First, for each of the top 250 most common inpatient procedures, we computed the *relative bias*, which we defined as the ratio between the rate of the procedure as measured in the healthcare claims dataset divided by the ground truth rate. Second, for the same procedures, we computed the strength of the association between that procedure and SDOH, in order to determine if the procedure is more likely to occur in neighborhoods with higher or lower socioeconomic status. Third, we tested the association between the bias (from step 1) and the relationship with SDOH (from step 2). The analysis was conducted in Python version 3.8.5. The analysis code posted at github.com/alex-dahlen/ClaimsDataBenchmarking.

**Step 1: Computing the relative bias associated with using claims data**

Our primary analysis focused on the cohort of people 18-64 residing in 5 states: CA, IA, MA, MD, and NJ. Inpatient procedures were classified using the Clinical Classification Software Refined (CCRS) classification of ICD-10-PCS codes, which buckets the procedure codes into identifiable procedures.

The states we studied made slightly different decisions around which facilities are included in the SID data: for example, while all states include data from acute-care facilities, California also includes data from psychiatric facilities and rehab centers.  To account for this, we restricted our analysis to acute care facilities.  Specifically, for the SID data from CA, we focused on facilities which were tagged with HospitalUnit = 1, which corresponds to acute care facilities, and for the SID data from MD, we removed facilities tagged with Hospital Unit = 6, which corresponds to psychiatric facilities. For the MarketScan data, we only included inpatient visits that received at least one tag of a location / place-of-service code = 10, which corresponds to Inpatient Hospital. In addition, we excluded a small number of procedures which were highly disproportionately concentrated in a subset of the states; these procedures were identified by extreme outliers in the chi-square distribution (>10,000) and included: psychotherapy for mental health, pharmacotherapy for mental health, substance use detoxification, and isolation procedures.  The goal of this process was to focus on facilities that identify as acute-care facilities and procedures that primarily occur in acute-care facilities. We focused our study on the 250 most common inpatient procedures in the ground truth (SID) cohort.

The rate of each procedure was evaluated separately using the claims data and then again using the ground truth (SID and census) data. To determine the rate of the procedure in the claims data, we divided the total number of inpatient discharges with at least one of the relevant procedure codes in MarketScan by the total number of patient-years of coverage in MarketScan, which was 2,325,710.9 person-years for our cohort. To determine the ground-truth rate of the procedure, we divided the count of inpatient discharges with at least one of the relevant procedure codes in SID by the total number of people age 18-64 in the census, which was 42,062,788. (Note: this assumes that the census tally of the total population 18-64 in the 5 states was stable over the year.)

We defined the relative bias for each procedure as the ratio of the MarketScan prevalence estimate divided by the SID prevalence estimate:

$$Relative bias = \frac{Claims data estimate}{Ground truth estimate} .$$

If the relative bias = 1, that indicates no bias (MarketScan-derived estimates align with the SID-derived estimate). Values less than 1 indicate that MarketScan underestimates the prevalence of a specific inpatient procedure, and values greater than 1 indicate that MarketScan overestimates the prevalence of a specific inpatient procedure. A histogram of the relative biases for the top 250 procedures is shown in **Supp. Fig. 1**, and a forest plot of the same - information is shown in Fig. 1. A table of the rates in the claims data and in the ground truth data, as well as the corresponding bias for all 250 procedures is provided in Supp. Table 1. The 95% Poisson confidence intervals were computed for both rate estimates, and were propagated to the bias estimate using the delta method for the log of the ratio.

**Step 2: Computing the strength of the association between procedure rate and SDOH**

This step required selecting a zip code-level proxy metric for SDOH. Our primary analysis used National Deprivation Index (NDI) as this proxy metric. NDI is defined by the National Cancer Institute, which combines 13 socioeconomic indicators drawn from the 2017 census. It is defined at the census tract-level, and we used population-weighted averages to roll it up to the zip code-level.

As a sensitivity analysis, we also defined a single metric to measure the SES of a zip code, and then we used a negative binomial model to measure the association between that SES metric and the procedure rate. To generate this single SES metric, we used Principal Component Analysis (PCA) to dimensionally reduce 25 different socioeconomic and demographic indicators into a single feature. The 25 socio-economic features were extracted from the 2019 5-year ACS survey at the census tract level for all 50 states, and rolled up to the zip code-level using population weighting. (We used the HUDS USPS 2019 crosswalk files to do this roll-up.) These features were: total population; fraction of the population who identified as non-Hispanic Asian, non-Hispanic Black, Hispanic, non-Hispanic Other, and non-Hispanic White; fraction of the population in the age brackets 0-17, 18-39, 40-64, and 65+; fraction of households making yearly incomes of <$20k, $20k – $40k, $40k – $75k, $75k – $125k, $125k - $200k, and $200k+; fraction of the labor force that is unemployed; fraction of the population 18-64 that is uninsured; fraction of the population 25+ with education less than a high school, high school, some college, college, and graduate school; fraction of houses that are owner-occupied; median house price; and log base 10 of the population density in people per square mile. (Square mileage, which was used to determine population density, was taken from the 2019 Census Gazeteer files.). These sociodemographic features are the same as the ones we used in our previous paper.

We standardized these 25 zip code-level indicators and then used PCA to dimensionally reduce to a single feature, which explained 24.9% of the total variance. **Supp. Fig 2** shows how this single vector decomposes along the 25 original socioeconomic indicators; the indicator mostly tracks zip codes with larger household incomes and greater education. **Supp. Fig. 3** shows a map of this parameter across zip codes in the lower 48 states; cities and coastal regions of the Northeast and West have disproportionately higher values of this SES parameter.

Next, we computed the zip code-level association between the rate of each inpatient procedure and the SDOD proxy metric. To determine this association, we used a Poisson model of the form:

$$log(E[Y_{i}^{k}])= \beta_{0}^{k}+\beta_{1}^{k}\times\mathrm{NDI}_{i}+\log\left( \mathrm{pop}_{i} \right),$$

where *k* indexes the top 250 procedures, and *i* indexes the 16,612 zip codes in our SID data; $Y_{i}^{k}$ is the count of the number of times the procedure occurred in zip code $i$, $\mathrm{NDI}_{i}$ is the value of the Neighborhood Deprivation Index in zip code $i$, and $\mathrm{pop}_{i}$ is the total population of that zip code, which serves as an offset for the model. We used the slope parameter $\beta_{1}^{k}$ as our measure of the strength of the association, since it measures the change in the log of the rate as you increase the NDI of a zip code by 1 unit. (For our sensitivity analysis, we repeated these models using our zip code-level SES proxy metric instead of NDI.)

The histogram of values of $\beta_{1}^{k}$ is shown in **Supp. Fig. 4** for both SDOH proxy metrics. In our primary analysis that uses NDI, A positive value of this parameter means that the procedure disproportionately occurs in zip codes with higher deprivation, and vice versa.

**Step 3: Using step 2 to explain the variation in relative bias**

Finally, we determined the procedure-level association between the relative bias (determined in step 1) and $\beta_{1}^{k}$ (determined in step 2), as shown in Fig. 2. We found that procedures that disproportionately occur in zip codes with low NDI are the ones that are most likely to be under-estimated ($p<0.0001).$To determine the extent of the association, we used a simple linear regression over the top 250 procedures, between a procedure’s bias in the claims dataset, and $\beta_{1}^{k}$, which measures the strength of the association between the procedure rate and SES. The only two terms in the regression were a slope and intercept, and the procedures were weighted by their ground truth rate. We reported the $R^{2}$ and the regression parameters in **Fig. 2**, and the results of our sensitivity analysis that uses our new SES variable in **Supp. Fig.**

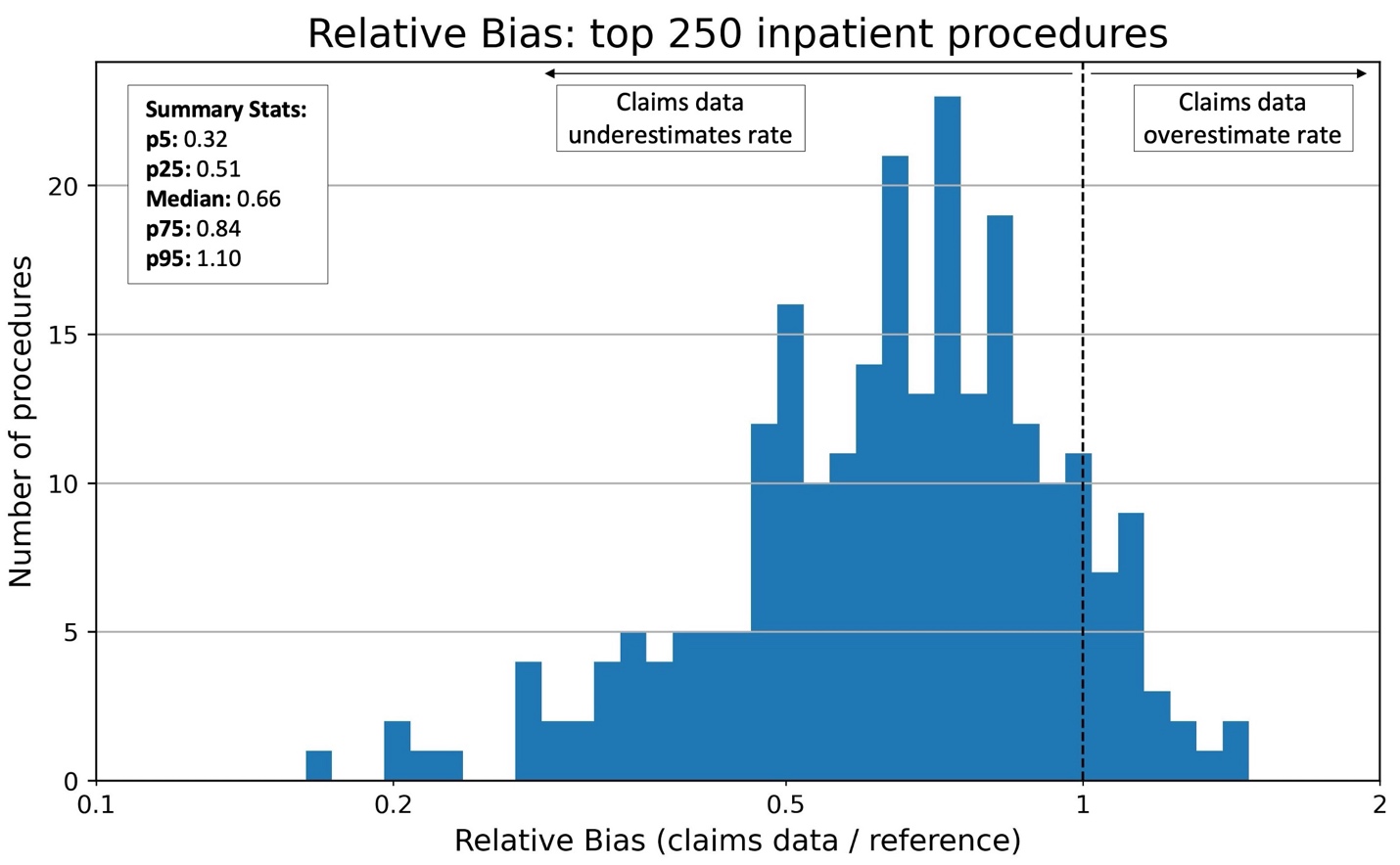

**Supp. Fig. 1.** A histogram showing the distribution of relative biases for the top 250 inpatient procedures. The relative bias is defined as the rate of a procedure computed in the healthcare claims data divided by the ground truth rate, so a relative bias of 1 would correspond to getting a “correct” answer in the claims dataset. This information is shown as a forest plot in **Fig. 1**.

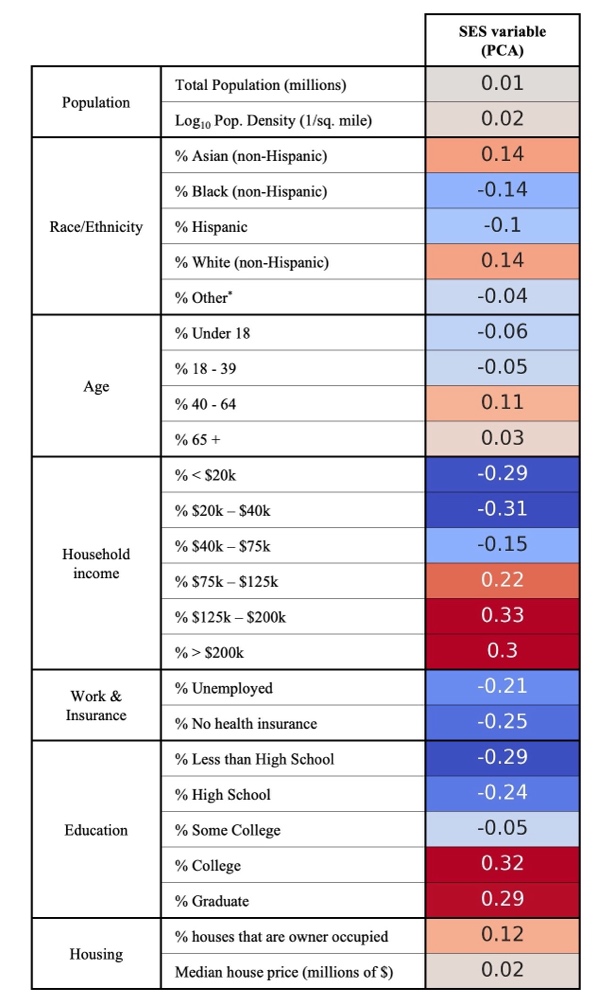

**Supp. Fig. 2.** We used PCA to dimensionally reduce 25 socioeconomic and demographic indicators, collected at the zip code level for all 50 states, down to a single SES parameter that explains 24.9% of the variance. This table shows how this single SES parameter decomposes along the 25 original indicators; it mostly tracks zip codes with larger household incomes and greater education.

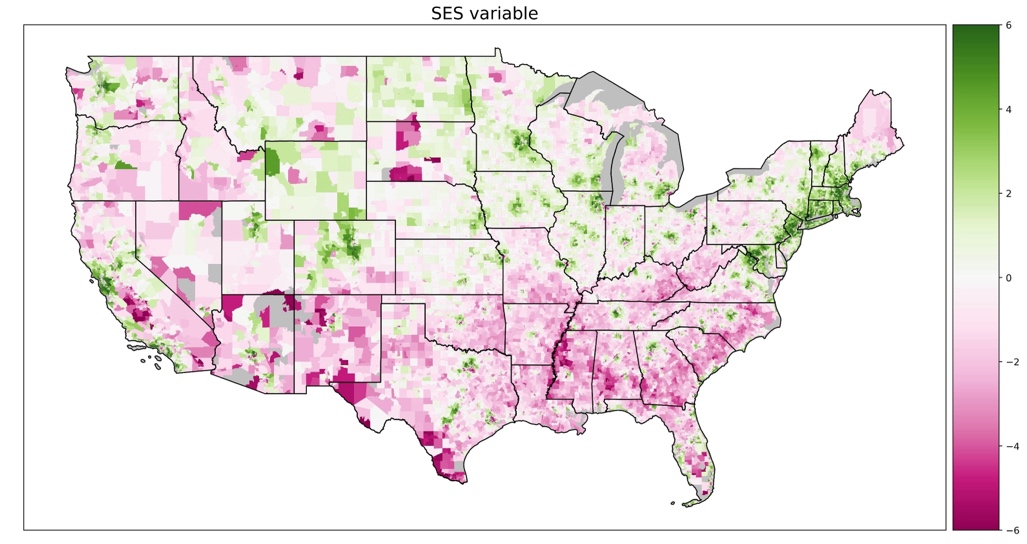

**Supp. Fig. 3.** A map of the value of the SES score for zip codes in the contiguous 48 states. The SES score is higher in cities and in coastal areas, and lower in rural regions and through the South.

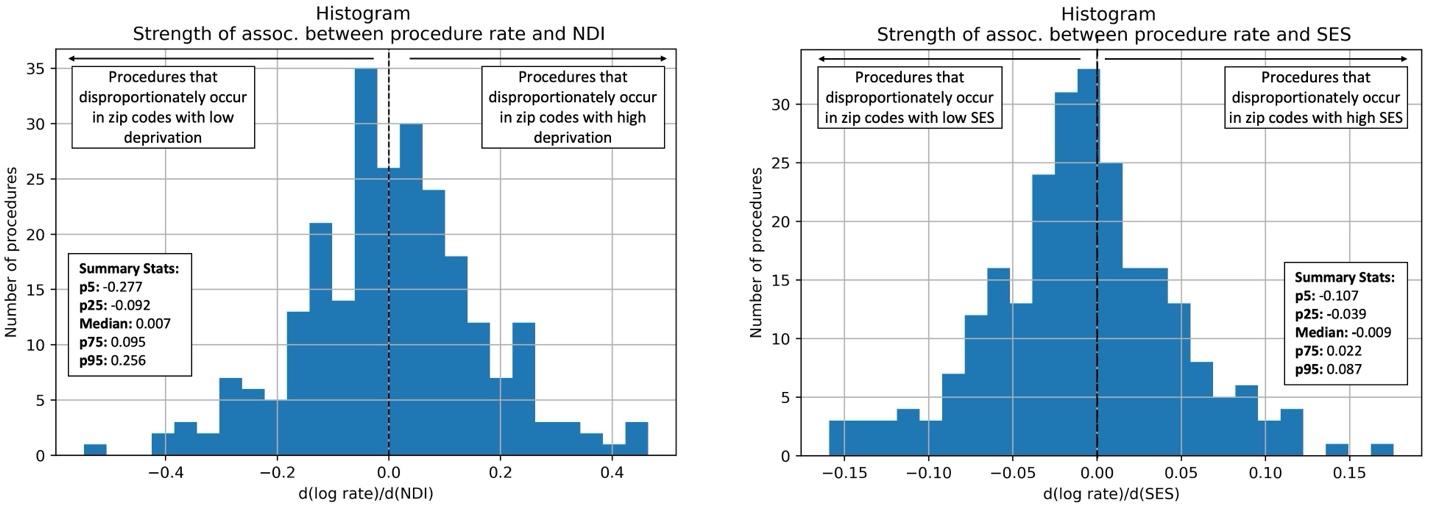

**Supp. Fig. 4.** For each of the 250 most common inpatient procedures, we generated a score to measure the strength of the association between the procedure rate and SES score. SES score is defined via our PCA procedure and the strength of the association is measured by a negative binomial model. Procedures with larger values of this score disproportionately occur in zip codes with high SES and vice versa.

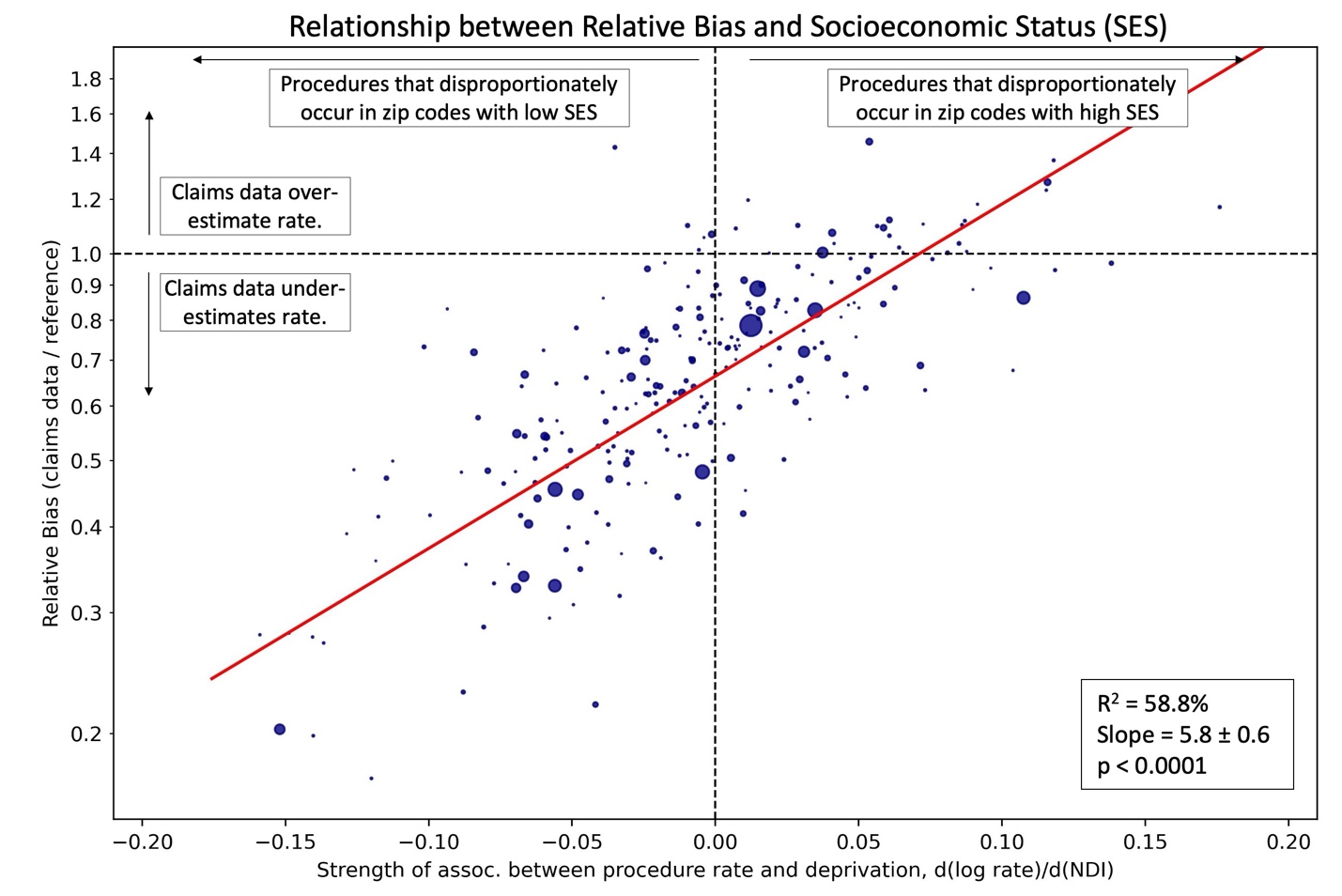

**Supp. Fig. 5.** A version of **Fig. 2** which uses our socioeconomic status metric (which is defined using 2019 census data) instead of neighborhood deprivation index (which uses 2017 data). We find an even stronger association than in Fig. 2, perhaps because of the difference in time periods. This figure shows a strong relationship where procedures that are disproportionately performed in zip codes with high SES are more likely to be overestimated in the claims data and vice versa.

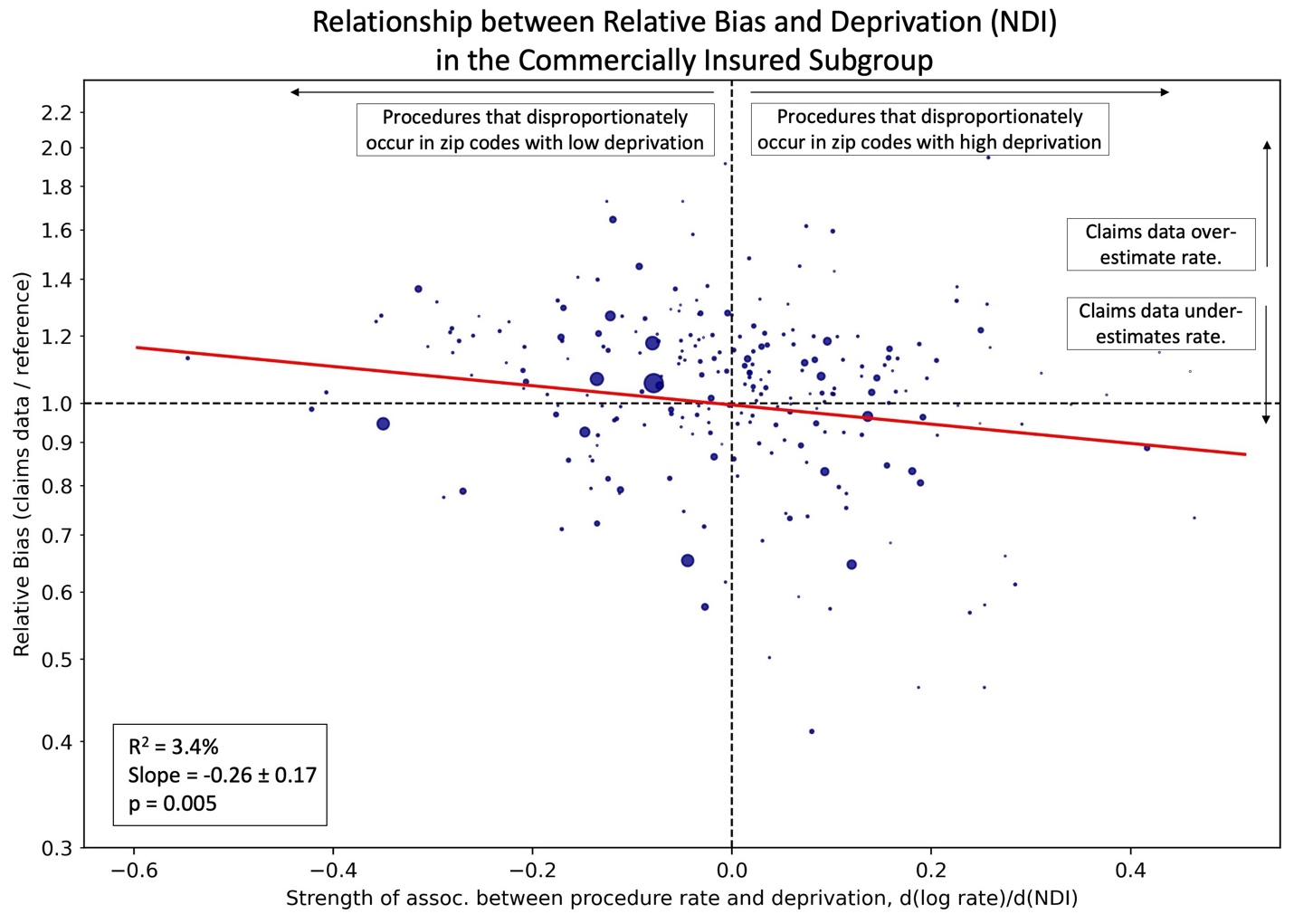

**Supp Fig 6.** The relationship between the relative bias (y-axis) for a procedure with respect to the commercially insured subgroup, and the strength of the association between each procedure and SDOH (x-axis), as measured by NDI. This figure is analogous to **Fig. 2**, except that here the relative bias for each procedure is measured compared to the commercially insured target population, while in **Fig. 2**, it is measured compared to the overall target population. The association is considerably smaller than in **Fig. 2**.

|  | **Main Cohort** | **Insured**  **Subgroup** | **Commercially Insured Subgroup** |
| --- | --- | --- | --- |
| **% significantly biased** | 93.2 | 93.6 | 76.4 |
| **% underestimated by factor of ≥2** | 22.8 | 24.8 | 1.2 |
| **% underestimated by factor of ≥1.9** | 28.4 | 29.2 | 1.6 |
| **% underestimated by factor of ≥1.8** | 31.6 | 33.6 | 1.6 |
| **% underestimated by factor of ≥1.7** | 36.0 | 39.6 | 3.2 |
| **% underestimated by factor of ≥1.6** | 41.6 | 46.0 | 4.4 |
| **% underestimated by factor of ≥1.5** | 50.4 | 52.8 | 5.6 |
| **% underestimated by factor of ≥1.4** | 55.2 | 62.0 | 6.8 |
| **% underestimated by factor of ≥1.3** | 66.8 | 69.2 | 10.0 |
| **% underestimated by factor of ≥1.2** | 74.4 | 78.4 | 15.2 |
| **% underestimated by factor of ≥1.1** | 82.4 | 84.8 | 20.8 |
| **% overestimated by factor of ≥1.1** | 4.8 | 3.2 | 42.8 |
| **% overestimated by factor of ≥1.2** | 2.0 | 1.6 | 22.4 |
| **% overestimated by factor of ≥1.3** | 1.2 | 0.8 | 10.8 |
| **% overestimated by factor of ≥1.4** | 0.8 | 0.4 | 5.6 |
| **% overestimated by factor of ≥1.5** | 0 | 0 | 3.6 |
| **% overestimated by factor of ≥1.6** | 0 | 0 | 2.8 |
| **% overestimated by factor of ≥1.7** | 0 | 0 | 2.0 |
| **% overestimated by factor of ≥1.8** | 0 | 0 | 1.2 |
| **% overestimated by factor of ≥1.9** | 0 | 0 | 1.2 |
| **% overestimated by factor of ≥2** | 0 | 0 | 0.4 |

**Supp. Table 1.** Percent of the 250 procedures we studied that are under- and over-estimated by factors of 1.1-2.0, in the three target populations of interest (columns). The top row shows the % of procedures that are statistically significantly biased at the 5% level: because of our large sample size, confidence intervals are very narrow.
